# Non-linear Measures of Gait Adaptability in Multiple Sclerosis - Sensitivity and Neurological Correlates

**DOI:** 10.1101/2022.07.06.22277289

**Authors:** MG Panisset, LE Cofré Lizama, L Peng, MP Galea

## Abstract

Multiple sclerosis (MS) is the leading cause of non-traumatic disability among young and middle-aged adults. People with MS (pwMS) rate walking and mobility as their highest priority for both research and symptom management. Given the importance of early initiation of disease-modifying therapeutics (DMTs) to minimize long-term disability, tools to identify early disease activity are needed. Traditional measures of disability, the Expanded Disability Status Scale (EDSS) and gait speed tests demonstrate poor reliability and responsiveness in cases with minimal disability. Nonlinear measures of gait, Local dynamic stability (LDS), Complexity (sample entropy) and Regularity (autocorrelation), measured in laboratory settings is sensitive to subclinical gait deterioration in people with MS (pwMS). These measures have not been tested in a clinical setting using wearable sensors.

**Methods:** Gait metrics were calculated in MatLab from inertial data collected from 59 pwMS (EDSS 0-4) and 23 age- and sex-matched healthy controls (HC) during a 5-minute walk. We aimed to provide known-groups validation of non-linear gait measures and compare sensitivity of LDS from sensors placed at sternum and sacrum in pwMS (ROC analysis). Associations of gait metrics with disability, Kurtzke Functional System scores and 3T MRI segmental brain volumes were examined.

**Results:** Most sternum-derived LDS measures detected significant differences between HC and pwMS (EDSS0-1) with moderate to large effect sizes (η^2^=.100-305), while the effect sizes for sacrum-derived LDS were lower (η^2^=.104-.166). Sternum_3D_ best distinuished EDSS_0-1_ from HC whereas the effect size was lower for gait speed (η^2^=.104). Sternum Instability-3D showed strongest correlation with pyramidal dysfunction (r_s_=.455, p<.001). Sensory dysfunction correlated significantly with Regularity in the vertical plane from both sensors, while cerebellar dysfunction was significantly associatesd with sacrum Regularity in the saggital plance (r=-.343, p=.008) and brainstem dysfunction with Complexity in the frontal plane (r=-.343, p=.008).

**Conclusion:** Sternum-derived LDS measures were more sensitive than Sacrum-derived measures. Correlations with clinical and morphological brain measures support the validity of walking deterioration as reflective of neurodegeneration in subcortical grey matter. The current findings of high sensitivity in non-disabled cases, as well as the clinical feasibility and relatively low costs, support the utility of these measures as a supplementary clinical assessment tool.

## 1. Introduction

Multiple sclerosis (MS) is the leading cause of non-traumatic disability among young and middle-aged adults. Estimated global prevalence is 2.2 million^1^ and over 33% of people with MS (pwMS) report disability related to mobility problems, resulting in loss of independence and reduced quality of life.^2^ The Expanded Disability Status Scale (EDSS), the standard metric of MS disability, demonstrates poor reliability and responsiveness in cases with minimal disability.^3^ Likewise, the timed 25-foot walk test (T25fwt), the traditional measure of gait deterioration related to disability, also has shown limited utility in early stages, failing to detect patient-reported mobility impairment in people with an EDSS score <2.0. ^4^

Barriers to accessing MRI, such as availability and costs, can lead to diagnostic delay in MS. Given the importance of timely initiation of disease-modifying therapeutics (DMTs) to minimize long-term disability, readily available and cost effective tools to identify early disease activity are needed.^5^ Emerging evidence suggests that gait metrics obtained from wearable sensors may provide reliable, sensitive and clinically-relevant digital biomarkers for disease progression in neurological conditions, including MS.^6, 7^ However, commonly reported spatiotemporal changes in gait seen in pwMS with moderate or greater disability, such as shorter step length and longer stance time,^8^ are not sensitive to subtle gait impairments in early MS.^9^ Emerging evidence suggests that non-linear metrics, such as the local dynamic stability exponent (LDS), or Lyapunov exponent, may be a sensitive marker of gait deterioration in early stage MS.^6, 10^

LDS measures the ability of the locomotor system to accommodate small step-to-step perturbations occurring from environmental (uneven floor surfaces) or internal sources (neuro-control or prediction errors).^11^ LDS has been shown to discriminate between minimally-affected pwMS (EDSS=1.2±0.9) and healthy participants (HC),^6^ predict future falls in pwMS (EDSS 3-4),^12^ and demonstrate more sensitivity to change after physical rehabilitation than gait speed (median EDSS 5).^13^

While investigations examining LDS to date have been performed under controlled conditions in laboratory settings,^14, 15^ clinical feasibility has not yet been examined. Furthermore, these examinations were conducted on a treadmill, which may mask naturally occurring gait pattern fluctuations.^14, 15^ Thus, we sought to examine the discriminative validity of LDS measured using accelerometers during overground walking in a clinical setting. Additionally, we sought to compare sensitivity of the LDS exponent calculated from two sensor locations: sacrum, which is the most commonly examined location, and sternum.^16^ Although placement of an inertial sensor on the upper trunk is less common, upper trunk movements may be more sensitive to MS-related motor impairments.^10^ Further, we aimed to explore the utility of two other non-linear measures of gait, sample entropy as a measure of complexity and autocorrelation as a measure of regularity. Lastly, we examined the neurological correlates of these measures using regional morphological volumes from coincident clinical 3T MRI.

The main aim of this study was to determine differences in local dynamic gait stability, complexity and regularity between non-disabled to moderately-disabled pwMS (EDSS 0-4) and healthy controls (HC) using inertial sensors in a clinical environment. Secondary aims were to: a) assess the discriminative validity and sensitivity of LDS measures by comparing non-disabled pwMS (EDSS_0-1_) and HC, b) compare the relative sensitivity of different sensor placements and c) explore construct validity by examining the relationship with EDSS, Kurtzke functional system (KFS) scores and morphological (volume) measures from clinical brain MRI.^3^ We hypothesised a) that pwMS would be less dynamically stable (greater LDS), less complex (lower sample entropy) and less regular (lower autocorrelation) than HC, even in the absence of evident gait disability, b) that sacral sensor would provide more sensitive measures, and c) that LDS would be positively associated with EDSS, pyramidal and cerebellar KFS scores, and negatively associated with normalised cortical and subcortical grey matter volumes.^17, 18^

## Methods

### Participants

The study was approved by the Human Research Ethics Committee at the Royal Melbourne Hospital (HREC2019.093). Sixty adults with relapsing-remitting MS (EDSS 0-4.0) with a diagnosis duration <15 years were recruited from the MS Centre at RMH from September 2019 to May 2022. Twenty three age- and sex-matched healthy adults were recruited from January to April 2021. Exclusion criteria were: other conditions affecting mobility, unable to follow instructions, pregnancy >5 months, or <5 months post-partum.^19^ For pwMS, additional exclusion criteria were recent changes in DMT (<4weeks), or unable to provide minimum data. Participants provided written informed consent. Previously, a sample size of 30 was deemed sufficient to detect group differences with using LDE from motion capture data (more sensitive) during treadmill walking (less sensitive).^6, 15^ We chose a feasible, roughly double sample size to accommodate for potential differences in sensitivity compared to previous reports.

### 2.4 Procedure

Participants walked at self-selected speed for 5 minutes along a 20-meter walkway in a lesser-used hospital corridor, given standardised instructions (Appendix A).^11^ Six sensors (Opals-APDM, Portland, Oregon, USA) were placed on the a) sacrum, b) sternum just below the sternal notch, c) wrists and d) feet, dorsally, just below the talocrural joint, over the shoes.

### 2.5 Descriptive Measures

Age, sex, height, weight, shoe type, self-reported exercise frequency, and rate of perceived exertion were recorded. Coincident EDSS, Kurtzke functional system scores (KFS) and morphological values were extracted from the local MS research database and digital medical record. Gait speed was calculated by the APDM software as mean speed after omitting turns, inclusive of two steps before and after.

### 2.6 Gait Measures

#### Gait Speed

LDS was calculated for 3D accelerations and for each motion plane (vertical, mediolateral, and anteroposterior) for both sensors, sternum and sacrum (Sternum_3D,_ Sternum_VT,_ Sternum_ML,_ Sternum_AP,_ Sacrum_3D_, Sacrum_VT,_ Sacrum_ML_ and Sacrum_AP,_ respectively) (Appendix B). Briefly, 150 strides were extracted using heel contacts detected from the foot sensors. As dynamic stability measures the relationship between cycles of repetitive motion, turns, including two seconds before and after peak rotational acceleration, were omitted.^20, 21^ Data from each lap were concatenated and normalized to 150 strides*100 data points, as described previously.^20, 22^ The LDS measures the divergence rate of a signal (i.e. trunk acceleration) as a reflection of small perturbations occurring during walking; the larger the LDE value, the less stable the walking pattern. LDS was calculated using the median dimensions (m) and time delay (t) for each plane and sensor across all trials, using the Rosenstein’s method in the 0-0.5 stride region.^23^ LDS-3D was calculated using a fixed m of 9 and t of 10.^6^ Sample entropy over accelerations signals measures the Complexity of the neuromuscular system’s behaviour by quantifying the probability that neighbouring points in the acceleration time-series will be within a predetermined range; values that are too low and too high can indicate a system is too chaotic or too rigid. To quantify the Regularity of walking, we calculate the autocorrelation; low values indicate a walking pattern that is highly variable, whereas values of 1 indicate very predictable and less adaptable system.

### 2.6 MRI Analysis

The electronic medical records of all participants with MS were searched to identify MRI results available within 2 months of the gait assessment date. Values for Normalized measures of volumes for whole brain, total gray matter, total white matter, and segmented brain structures, obtained from clinical (3D) T1-weighted magnetisation prepared rapid gradient echo (MPRAGE) using an automated process (Morphobox), were extracted.^24^

### 2.7 Statistical Analysis

Statistical analyses were conducted in IBM-SPSS 27 (IBM Inc., Chicago, IL, USA). Normality of data distributions were assessed using the Shapiro-Wilkes test. Independent t-tests or Mann–Whitney U tests (if not normally distributed) were used to compare between-group differences in subject characteristics. The discriminatory capacity of gait measures was evaluated using the area under the Receiver Operating Characteristics curve (AUC). The AUC ranges from 0 to 1.0, with values of 1.0 for a perfect diagnostic test, and 0.5 or less for a useless test. AUC values 0.7-0.8 were considered fair, 0.80-0.90 were considered good, and any higher were considered excellent.^25^ Since a broad spectrum of case and control are required to evaluate the accuracy of specificity, from which the ROC is derived, the secondary analysis comparing only non-disabled pwMS (EDSS 0-1) to HC utilized ANCOVA, to allow controlling for confounding variables and estimations of effect size (Eta squared). Effect sizes (η^2^) were interpreted as small (.01), moderate (.06) and large (.14).^26^ Pearson’s correlations or Spearman’s (if non-normally distributed according to Shapiro-Wilkes test) explored relationships between sensor metrics and clinical variables. Correlations <.30 were deemed small, .30-.49 as moderate, and .50 and higher as large in terms of magnitude of effect sizes.^27^ All *p*-values are two-tailed. Significance was set at *p*<.05.

### 2. Results

No adverse events occurred during testing. Clinical characteristics of all pwMS and each disability group are presented in Table 2. There were no significant differences in sex, age, or height between HC and pwMS (Table 1). However, pwMS had a significantly higher body mass index (27.6(5.0) versus 24.0(4.3) for HC).

**Table 1.** Participant Characteristics

|  | Healthy Controls |  | All PwMS (n=59) |  | EDSS 0-1 (n=20) |  |
| --- | --- | --- | --- | --- | --- | --- |
|  | mean | SD | mean | SD | mean | SD |
| Age | 44.3 | 11.1 | 42.6 | 10.9 | 37.5 | 8.8 |
| Sex (% male) | 30% |  | 32% |  | 50% |  |
| Height (cm) | 171.5 | 8.7 | 170.7 | 9.5 | 173.1 | 10.4 |
| Body Mass Index (kg/m<sup>2</sup>) | 24.0 | 4.3 | 27.6* | 5.0 | 26.6 | 6.3 |

Abbreviations: PwMS, people with multiple sclerosis; EDSS extended disability status scale; SD, standard deviation. \* Statistically different from healthy controls Mann-Whitney U=349, p=.0001

**Table 2.**
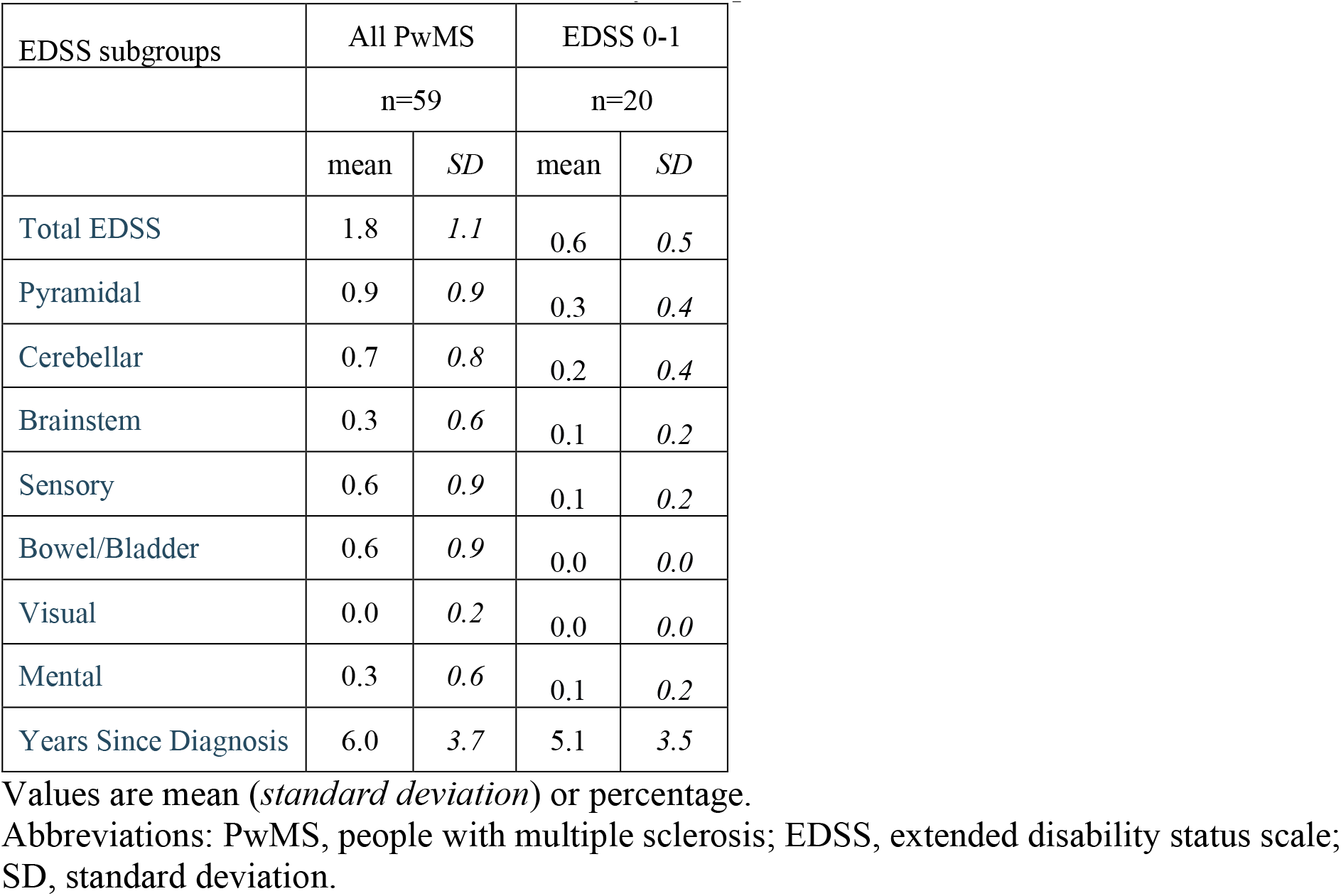
Clinical Characteristics of Disability Groups

| EDSS subgroups | All PwMS |  | EDSS 0-1 |  |
| --- | --- | --- | --- | --- |
|  | n=59 |  | n=20 |  |
|  | mean | SD | mean | SD |
| Total EDSS | 1.8 | 1.1 | 0.6 | 0.5 |
| Pyramidal | 0.9 | 0.9 | 0.3 | 0.4 |
| Cerebellar | 0.7 | 0.8 | 0.2 | 0.4 |
| Brainstem | 0.3 | 0.6 | 0.1 | 0.2 |
| Sensory | 0.6 | 0.9 | 0.1 | 0.2 |
| Bowel/Bladder | 0.6 | 0.9 | 0.0 | 0.0 |
| Visual | 0.0 | 0.2 | 0.0 | 0.0 |
| Mental | 0.3 | 0.6 | 0.1 | 0.2 |
| Years Since Diagnosis | 6.0 | 3.7 | 5.1 | 3.5 |

Values are mean (*standard deviation*) or percentage.
Abbreviations: PwMS, people with multiple sclerosis; EDSS, extended disability status scale; SD, standard deviation.

Age showed weak but significant association with only one gait measure, Mediolateral Step Regularity (r_s_= .260, p= .019), which was also correlated with years since diagnosis (.374, p=.004) and BMI (rs=.293,p=.043). BMI correlated weakly to moderately with several gait measures from the sacral sensor, most strongly with Anteroposterior Complexity (r=.347, p=.002) and Mediolateral Stride Regularity (r=-.345, p=.002). Gait speed did not significantly correlate with age, disease duration or BMI, however there was a significant but weak association with some LDE measures, the strongest being Sternum3D (rs = .288, p=.009)

### 2.1 Classifying MS using Area Under the ROC curve

AUC values for all gait measures are presented in Table 3. With AUC <.7 (0.682) gait speed was found to have less than fair diagnostic power. SternumVT had fair accuracy, while the other LDE measures from the Sternum were good. None of the other measures demonstrated utility for classifying pwMS compared to HC.

**Table 3.** Analysis of Gait Measures

|  |  |
| --- | --- |
| Area Under the Curve | ANCOVA |

| Classifying pwMS (EDSS 0-4) from Healthy Controls |  |  |  |  |  | Comparing pwMS (EDSS 0-1) to Healthy Controls |  |  |
| --- | --- | --- | --- | --- | --- | --- | --- | --- |
| Sensor |  | 95% Confidence Interval |  |  |  |  |  |  |
| Measure | Plane | AUC | Lower Bound | Upper Bound | p | F | p | Eta Squared |
| **Sternum** |  |  |  |  |  |  |  |  |
| LDE | 3D | **0.810** | 0.711 | 0.908 | 0.000 | 13.039 | **<.001** | 0.261 |
|  | vertical | **0.766** | 0.653 | 0.880 | 0.000 | 16.216 | **<.001** | 0.305 |
|  | mediolateral | **0.805** | 0.684 | 0.925 | 0.000 | 4.124 | **0.050** | 0.100 |
|  | anteroposterior | **0.818** | 0.711 | 0.925 | 0.000 | 7.655 | **0.009** | 0.171 |
| Complexity | vertical | 0.571 | 0.435 | 0.707 | 0.329 | 0.354 | 0.555 | 0.009 |
|  | mediolateral | 0.384 | 0.249 | 0.518 | 0.111 | 1.237 | 0.273 | 0.032 |
|  | anteroposterior | 0.322 | 0.181 | 0.464 | 0.015 | 5.648 | **0.023** | 0.132 |
| Regularity - Step | vertical | 0.225 | 0.121 | 0.329 | 0.000 | 4.062 | 0.051 | 0.099 |
|  | mediolateral | 0.450 | 0.304 | 0.596 | 0.494 | 0.214 | 0.646 | 0.006 |
|  | anteroposterior | 0.439 | 0.297 | 0.581 | 0.404 | 0.599 | 0.444 | 0.016 |
| Regularity - Stride | vertical | 0.207 | 0.103 | 0.311 | 0.000 | 5.473 | **0.025** | 0.129 |
|  | mediolateral | 0.355 | 0.216 | 0.493 | 0.046 | 5.541 | **0.024** | 0.130 |
|  | anteroposterior | 0.417 | 0.271 | 0.563 | 0.255 | 0.037 | 0.848 | 0.001 |
| **Sacrum** |  |  |  |  |  |  |  |  |
| **Gait Speed** | 3D | 0.682 | 0.552 | 0.812 | 0.011 | 4.280 | **0.046** | 0.104 |
| LDE | 3D | 0.679 | 0.543 | 0.815 | 0.014 | 5.378 | **0.026** | 0.127 |
|  | vertical | 0.665 | 0.524 | 0.806 | 0.024 | 7.370 | **0.010** | 0.166 |
|  | mediolateral | 0.621 | 0.486 | 0.756 | 0.098 | 2.892 | 0.097 | 0.072 |
|  | anteroposterior | 0.591 | 0.450 | 0.731 | 0.214 | 5.094 | **0.030** | 0.121 |
| Complexity | vertical | 0.528 | 0.376 | 0.679 | 0.704 | 0.765 | 0.387 | 0.020 |
|  | mediolateral | 0.448 | 0.307 | 0.588 | 0.473 | 0.351 | 0.557 | 0.009 |
|  | anteroposterior | 0.621 | 0.474 | 0.769 | 0.096 | 0.495 | 0.486 | 0.013 |
| Regularity - Step | vertical | 0.263 | 0.148 | 0.379 | 0.001 | 2.509 | 0.122 | 0.064 |
|  | mediolateral | 0.344 | 0.218 | 0.469 | 0.032 | 2.021 | 0.164 | 0.052 |
|  | anteroposterior | 0.333 | 0.204 | 0.463 | 0.022 | 0.669 | 0.419 | 0.018 |
| Regularity - Stride | vertical | 0.257 | 0.144 | 0.371 | 0.001 | 3.643 | 0.064 | 0.090 |
|  | mediolateral | 0.279 | 0.158 | 0.399 | 0.002 | 3.112 | 0.086 | 0.078 |
|  | anteroposterior | 0.310 | 0.184 | 0.437 | 0.009 | 2.346 | 0.134 | 0.060 |

ANCOVA has controlled for age and BMI

### 2.2 Non-disabled pwMS

All LDE measures were able to distinguish between EDSS_0-1_ and HC with moderate to large effect sizes (η^2^=.1 to .3), except Lumbar_ML,_ when controlling for age and BMI (Table 3). Sacrum_VT_ and Sacrum_3D_ demonstrated the highest effect sizes (η^2^=.305 and .261, respectively). Complexity (AP) and Stride Regularity (VT and ML) from the Sternal sensor demonstrated moderate effect sizes (η^2^=.129 to .132). There were no between-group differences in Complexity or Regularity from the Sacral sensor after controlling for age and BMI.

### 2.3 Correlations with Functional Systems

Gait speed did not demonstrate a significant association with EDSS or any Kurtzke Functional Score. Total EDSS score correlated weakly to moderately with a number of non-linear gait measures, the strongest being from the Sternal sensor, with SternumML (r=.39, p=.002), Step Regularity (VT) (r=-.344, p=.008) and Complexity (VT)(.328, p=.011) all having moderate associations (Table 4). The highest correlation was between Sternum3D and pyramidal score (r=.455, p<.001), followed again by other sternal measures: SternumML (r=.384, p=.003), Step Regularity (VT) (rs=.361, p=.005), and Complexity (VT) (r=.348, p=.007). Cerebellar score was moderately associated with only Step Regularity (AP) from the Sacral sensor (r=-.343, p=.008); other associations were weak. Brainstem score showed a moderate association only with Sacral Complexity (ML) (rs=-.343, p=.008), no other associations with cerebellar score were significant. Sensory score showed moderate correlations in Step Regularity (VT) from the sternum (rs=-.351, p=.006) and sacral sensors (rs=-.327, p=.011). Visual system score weakly correlated with LumbarLDE_3D only. None of the gait measures were significantly related to Mental or Visual Functional Scores.

**Table 4.**
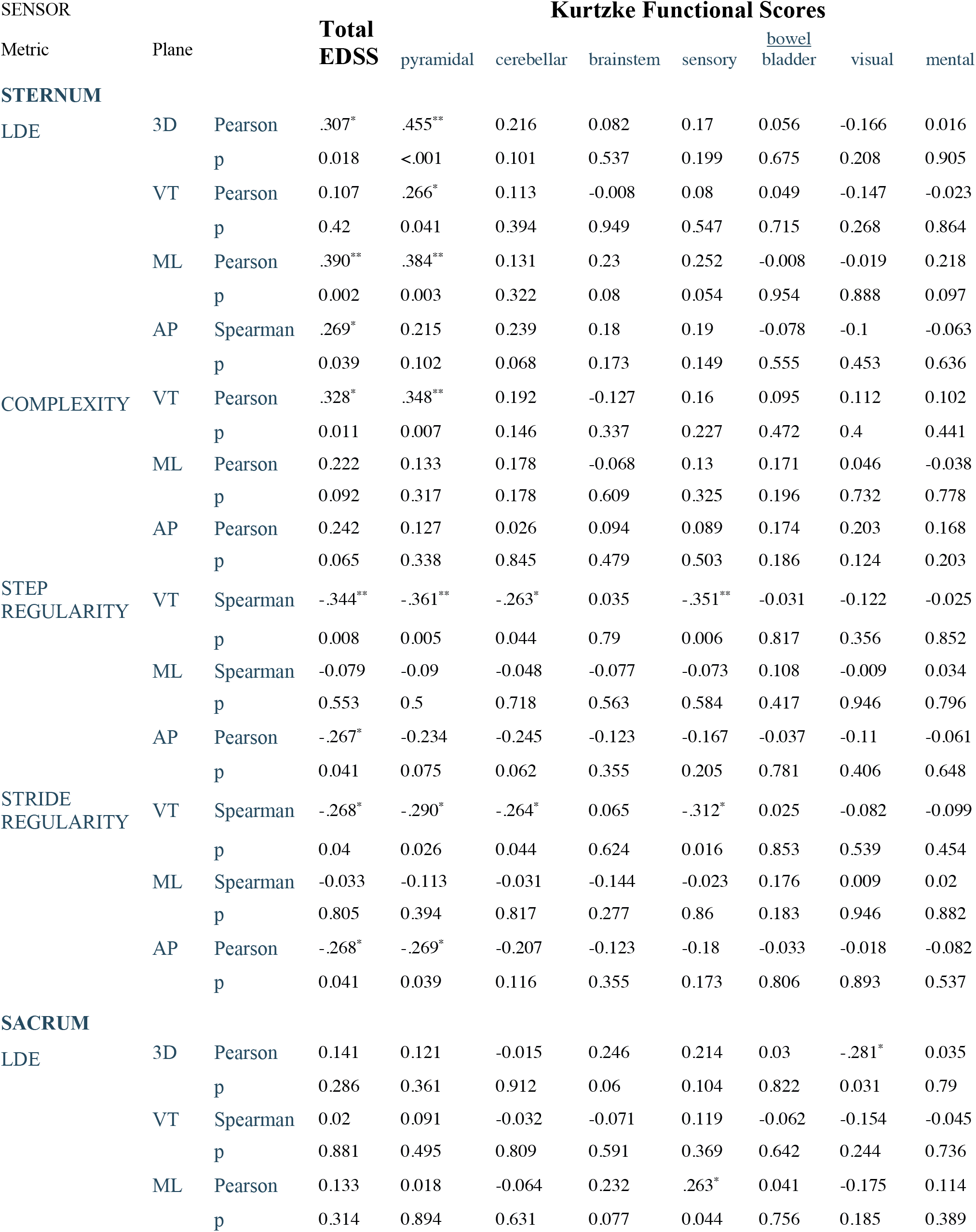
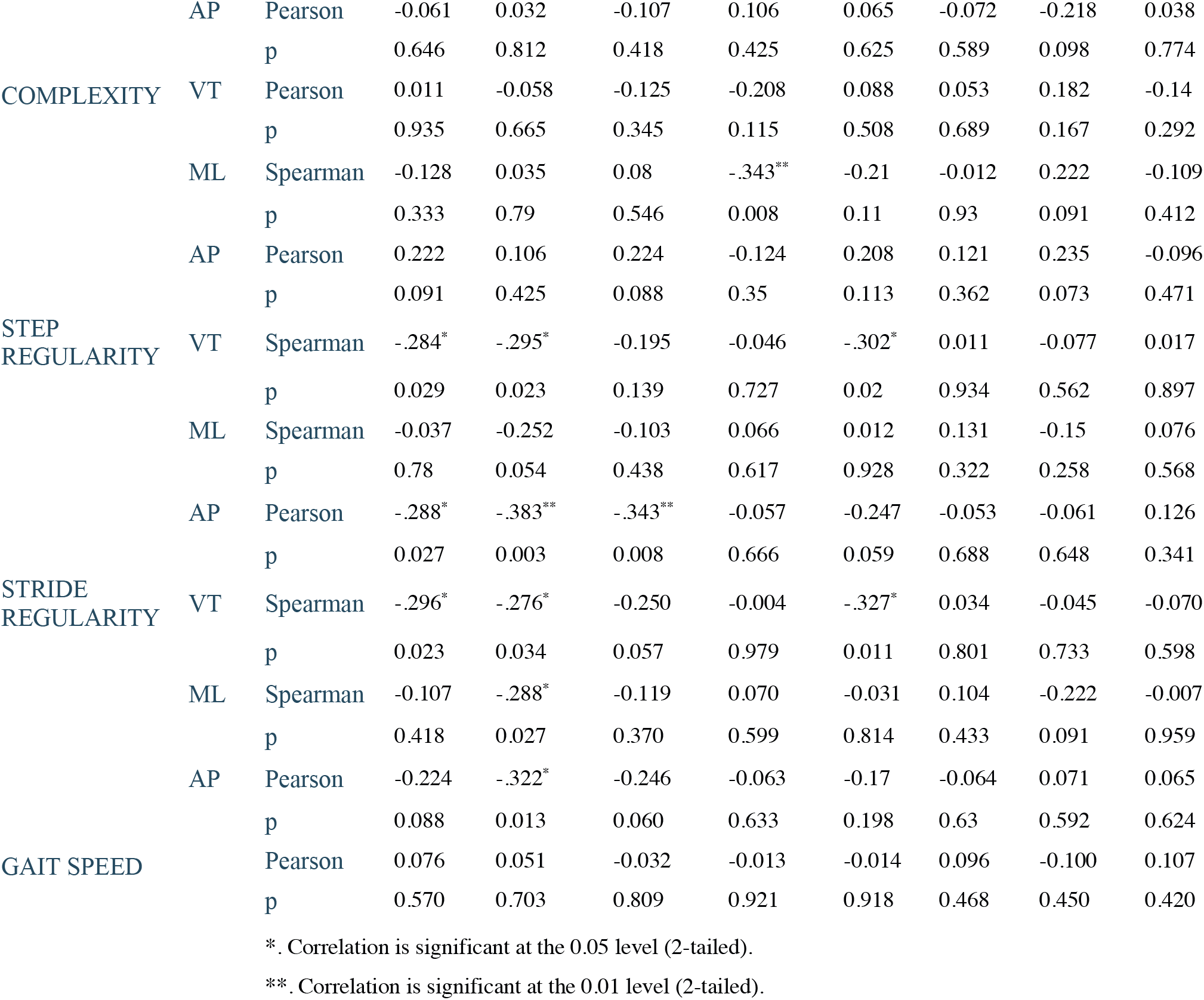
Correlations between non-linear gait metrics and Clinical Measures

### 2.4 Correlations with Brain Morphology

MRIs were available for 49 pwMS. Associations between gait measures and morphology measures were examined, controlling for age as a covariate (Supplemental Table A). Gait speed did not demonstrate a significant association with any morphology measure. The strongest association between a non-linear gait measure and segmental brain volume was between the right caudate and Sternum3D (r=-0.482, p<.001). The left thalamus was significantly associated with both Stride Regularity (AP) (r=0.432, p=.002) and Complexity (AP) (r=-0.394, p=.004) from the Sternum sensor. Stride Regularity (AP) also demonstrated a significant association with cerebellum volume (r=0.405, p=.003), pons volume (r=0.371, p=.007) and Mesencephalon volume (r=0.366, p=.008). Normalised mid-sagital area of the corpus callosum and SternumLDE (ML) were uniquely associated (r=-0.358, p=.01).

## 3. Discussion

The aim of this study was to test the discriminative validity of three non-linear measures of walking adaptability derived from data recorded during overground walking in a clinical setting using wearable inertial sensors at the sternum and sacrum, and examine their relationship to common clinical measures. Our results of lower dynamic stability in pwMS compared to HC are consistent with previous laboratory studies.^6, 12, 21, 28, 29^ As expected, AUC analysis revealed that LDS from the Sternum showed greater diagnostic utility than gait speed when EDSS **≤**4. Further, we found, for the first time, that LDS was sensitive enough to discriminate between HC and non-disabled pwMS (EDSS_0-1_). Importantly, these findings are based on neurological diagnosis of MS, rather than disability score, as nine participants had normal neurological examination at the time of testing (EDSS=0). This highlights the potential diagnostic utility of these digital biomarkers early in asymptomatic MS, or for monitoring those at high risk of MS, such as family members.

Our finding that sternum LDS measures were more sensitive than sacral LDS measures in the broader EDSS_0-4_ group was unexpected. We hypothesized that sacral LDS would be more sensitive, given that the sensor approximates the body’s centre of mass, and hence would best represent overall dynamic stability.^28^ Our results are in line with recent evidence that, although LDS values are higher at the sacrum than the sternum in both pwMS and HC, pwMS (EDSS_0-5_, mean(SD) 2.0(1.1)) had comparatively higher LDS at the upper trunk when walking overground.^10^ We found that pyramidal KFS was most closely associated with sternum LDS measures, suggesting that the task of stabilizing the upper trunk might be responsive to physical rehabilitation.^13^ Interestingly, Peebles et al. found no correlation between any sternum LDS and pyramidal score during treadmill walking in a similarly disabled cohort.^30^ This highlights the need to compare similar protocols only, when interpreting the literature, as treadmill walking is known to mask gait instability to a considerable extent.^15^

The current findings suggest that the ability to stabilise the upper trunk and, to a lesser extent, the pelvis is impaired in sub-clinical MS. This may indicate that an impaired motor control system prioritizes a tighter control of dynamic stability closer to the body’s centre-of-mass (CoM), for example, to avoid falling. On one hand, the observed trunk instability could be indicative of effective compensatory adjustments aimed at maintaining stability of the CoM (sacrum LDS) when subclinical lower limb impairments such as ankle weakness and/or co-contraction may lead to gait instability.^31^ There is some evidence to support this hypothesis. Decreased activation of medial gastrocnemius has been found in early MS,^32^ and pwMS (median EDSS 3.5) demonstrated greater sagittal plane excursion of the trunk than HC during overground walking, with the effect being stronger during push-off of the weak side.^33^

On the other hand, dynamic instability of the upper trunk is likely to affect the ability to stabilize the head, and thereby the visual and vestibular systems, which are critical for balance control.^34^ Indeed, upper trunk LDS has been shown to predict future falls in pwMS (EDSS 3-4),^12^ and was associated with falls history in people with MS (mean EDSS 2.9 (1.2))^29^ and in older people,^35^ while sacrum LDS was unable to predict future falls in older people.^36^ In the next sections, we look at the performance of LDS metrics according to each plane of motion.

### 4.1 Frontal Plane

Sternum_ML_ demonstrated the highest effect size of all metrics when comparing HC with EDSS_0-1_, and was the only measure associated with corpus callosum cross sectional area, which involved in interlimb coordination. This finding is clinically relevant when considering that Sternum_ML_ during treadmill walking was higher in MS fallers (EDSS 2.9 (1.2)).^29^ Furthermore, another study found Sacrum_ML_ was responsive to inpatient rehabilitation for falls prevention in pwMS with mobility limitations.^13^

### 4.2 Sagittal Plane

Our finding that LDE_Sternum_AP_ was greater in pwMS than HC is also consistent with previous evidence.^29, 37^ Interestingly, although Sternal Complexity-AP and Stride Regularity -AP did not distinguish pwMS from HC, they both were significantly associated with decreased thalamus volume, which was previously shown to be a predictor of future cognitive decline. ^38^ We found that Stride Regularity-AP was also associated with decreased cerebellar volume, while Step Regularity AP was associated with neurologist rated pyramidal and cerebellar function, lending some convergent validity that walking adaptability in the sagittal plan is associated with subcortical neurological impairments.

### 4.3 Vertical Plane

Sternum_VT_, and to a lesser extent Sacrum_AP,_ demonstrated a larger effect size for between-group difference. Previously, patients with cerebellar lesions demonstrated the highest effect size for Sacrum_VT_ (Cohen’s d 1.34) compared to Sacrum_ML_ and Sacrum_AP_ (Cohen’s d 1.11 and 0.65, respectively).^39^ In a different study, Sternum_VT_ during treadmill walking at self-selected speed showed a moderate correlation with cerebellar KFS, as well as a weak but significant correlation with sensorimotor delay.^30^ Based on this, one might postulate that LDS in the vertical plane may reflect motor control impairments in the timing or coordination of antigravity musculature. This is supported in part by our finding of a weak but significant association of LDE_SacrumVT with decreased pons and medulla volumes and of LDE_SternumVT with atrophy in the caudate, which is associated with posture and accuracy of directed movement. While we found no correlation between cerebellar KFS score and Sacrum_VT_, this might be due to the comparatively low level of cerebellar impairment in our cohort. Looking beyond pwMS, this is supported by findings that patients with cerebellar ataxia (CA) exhibited instability of limb loading at heel strike associated with an abnormal ground reaction force in the vertical direction and impaired intersegmental coordination.^40^ This evidence, combined with a higher LDS observed in the vertical plane during gait in another CA cohort, suggests that the cerebellum plays a major role in dynamic stability of the upper trunk.^39^

Another interesting finding in the vertical plane was that Step, and to a lesser extent, Stride Regularity were negatively associated with neurologist assessed sensory dysfunction. This is consistant with evidence of the importance of the cerebellar-sensorimotor network in motor control.^41^

### 4.4 Clinical relevance

Although gait speed showed only fiar diagnostic power, it was able to distinguish between HC and non-disabled pwMS, although the effect size was smaller than non-linear gait metrics. This highlights the need for more sensitive measures to monitor the onset of mobility impairments in early and prodromal disease.^5^

We found half the non-linear measures correlated only weakly with EDSS. Given the known lack of sensitivity when EDSS<4, this was unsurprising.^42^ Indeed, EDSS previously showed no correlation with total lesion load on T2 MRI, while both 2-min walk test and dynamic balance testing showed weak correlations with lesion load in early MS (<5 years).^4^ Previous studies have found little to no correlation of dynamic gait stability at the sacrum (LDS) with patient-reported outcome (PRO) measures of static postural balance, fatigue or disease impact,^37^ suggesting that poor dynamic stability may reflect impairments in complementary neuromotor systems not captured by the EDSS <4 and/or not adequately characterised by existing PROs.^30, 37^

Studies using LDS to quantify gait stability in non-fallers and fallers pwMS have found poorer stability in the latter group^12^, which correlated with physiological impairments (e.g. vibration threshold)^30^. A previous study found that gait stability measured at the upper trunk was a predictor of falls in pwMS with moderate disability (EDSS 3-4)^12^, further longitudinal studies should monitor LDS in pwMS at early stages of the disease to determine whether LDS can predict falls risk earlier, when physical rehabilitation or other disease-modifying treatments could be more effective.^43^

Few intervention studies have used LDS as an outcome measure in pwMS. One study found LDS was more sensitive to change than gait speed in evaluating effectiveness of 3-weeks in-patient rehabilitation for falls prevention in pwMS (median EDSS=5).^13^ Further investigations are needed to examine the sensitivity of LDS to change and its potential as an outcome measure.

### 4.5 Generalisability and Limitations

A strength of the study is its generalisability due to testing within a clinical setting. The main difference between a laboratory setting and this clinical setting was the presence of other people and conversations in the hallway and noise from the café at the end of the hallway, which would have provided intermittent distractions for all participants. We suggest this provided a better approximation of gait stability in daily life.

Variation in footwear between subjects might have affected outcomes. Studies in HC have shown that LDS from a foot or sacrum sensor was no different between barefoot walking versus shod.^44^ However, whether type of footwear effects LDS in pwMS is unknown. Another potential source of variability include seasonal thermosensitivity (n=8), however the tests were conducted in a climate-controlled hospital, minimising seasonal effects during testing. We had difficulty obtaining optimal placement of the sacral sensor in some participants due to small waist to hip ratio (n=5), however, the difference in placement was no greater than 5-10cm. Another limitation is the use of stitched acceleration time-series to calculate the LDS. However, this approach has been previously used to calculate LDS^10^ and other metrics^20^, and allows for implementation of overground testing in a clinical corridor.

## 4. Conclusion

Given the importance of early initiation of DMTs to minimize long-term disability, and challenges with diagnostic delay given the current diagnostic algorithm, more sensitive tools that can identify early signs of gait impairments are needed.^5, 45^ To our knowledge, this is the first time that any wearable-sensor-based gait measure has demonstrated discriminative validity between HC and non-disabled pwMS (EDSS 0-1). The current findings of high sensitivity in non-disabled cases, as well as the clinical feasibility and relatively low cost, support the potential utility of these measures as a supplementary clinical diagnostic tool in early MS (EDSS 0-4).

## Supporting information

Supplemental File - Morphology

## Data Availability

All data produced in the present study are available upon reasonable request to the authors

## Abbreviations

LDS: Local Dynamic Stability
AP: Anteroposterior
ML: Mediolateral
VT: Vertical
CoM: Center of Mass

## Acknowledgements

*The authors acknowledge the participants for their involvement, the RMH MS Centre Research and Administrative staff for assistance with recruitment and scheduling, and the RMH MS Clinic neurologists for contribution to recruitment*.

Tomas Kalincik served on scientific advisory boards for BMS, Roche, Janssen, Sanofi Genzyme, Novartis, Merck and Biogen, steering committee for Brain Atrophy Initiative by Sanofi Genzyme, received conference travel support and/or speaker honoraria from WebMD Global, Eisai, Novartis, Biogen, Sanofi-Genzyme, Teva, BioCSL and Merck and received research or educational event support from Biogen, Novartis, Genzyme, Roche, Celgene and Merck.

The remaining Authors declare that there is no conflict of interest.

The data that support the findings of this study are available from the corresponding author, [MGP], upon reasonable request.

---

**Figure 1.**
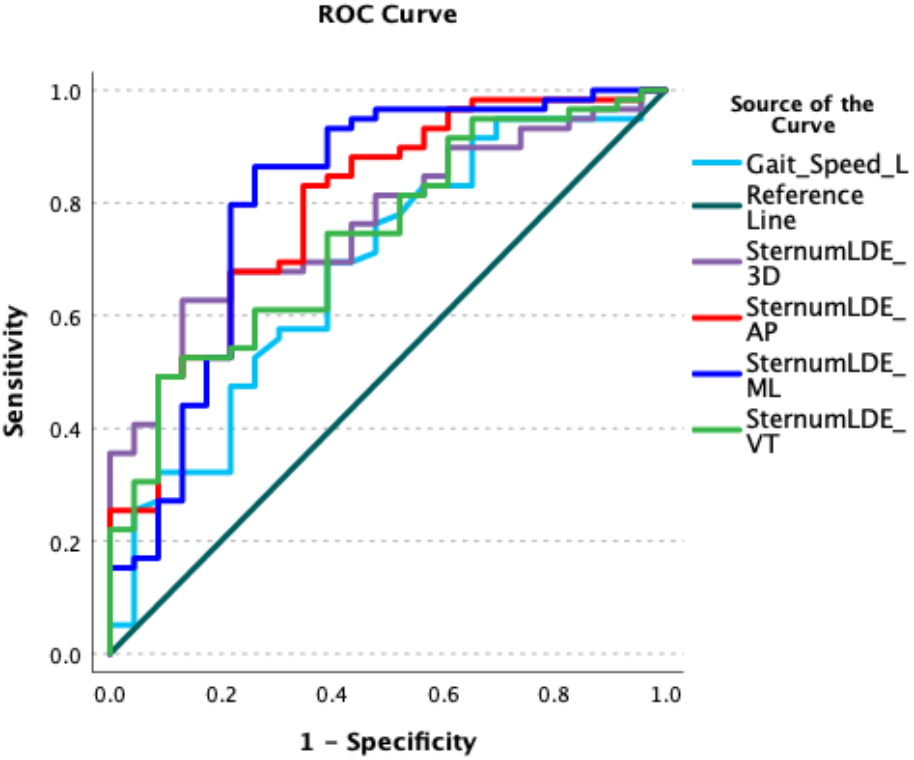
Reciever Operating Characteristic Curves for each variable using all data (EDSS 0-4)

